# Cholera rapid diagnostic tests at the host-microbe interface: Key Considerations for Global Deployments

**DOI:** 10.1101/2025.05.14.25327654

**Authors:** S. M. Ahmed, Md. Abu Sayeed, I. Sriguha, E. T. Cato, A. Creasy-Marrazzo, K. Islam, Md. I.UL. Khabir, Md. T. R. Bhuiyan, Y. Begum, Md. Taufiqul Islam, Zahid Hasan Khan, E. Freeman, A. Vustepalli, L. Brinkley, M. Kamat, L. S. Bailey, N. Madi, F. Qadri, B. J. Shapiro, D. T. Leung, K. B. Basso, D. A. Sack, J. R. Andrews, A. I. Khan, E. J. Nelson

## Abstract

**Background:** Effective cholera outbreak response requires accurate bedside rapid diagnostic tests (RDTs) because access to laboratories is often limited. Formative studies suggest cholera diagnostics have multiple vulnerabilities, including antibiotics and predation by bacteriophage (‘phage’) specific to *Vibrio cholerae* (*Vc*).

**Methods:** We conducted a prospective nationwide study in Bangladesh among over 2000 patients with diarrhoeal disease to characterize how these vulnerabilities impact RDT performance. Assays included culture, qPCR and mass spectrometry.

**Findings:** With the current gold standard of culture or qPCR *Vc* positivity, we found no effect of phage on RDT performance. When the diagnostic criteria were expanded to include phage, there was a small decrease in RDT sensitivity. In contrast, large increases in sensitivity and specificity were observed among patients with moderate and severe dehydration. Using the expanded definition, the odds of RDT positivity decreased among cholera patients with phage exposure. The effect was most robust among patients with severe dehydration. Antibiotic were detected in over 80% of samples by LC-MS/MS which limited testing for effects on RDTs. Applying these findings, we estimated that restricting RDT use to severe patients with no reported antibiotic exposure increases sensitivity by 50% compared to unrestricted use. If phage were a diagnostic proxy for *Vc*, we estimate RDT would miss an additional 17% of cholera cases.

**Interpretation:** Cholera RDTs have critical limitations that require consideration in global deployments. Inclusion of phage detection in diagnostic criteria may improve case detection which requires further study. The impact of these findings likely extends to other diseases where diagnostics share similar vulnerabilities.

**RESEARCH IN CONTEXT:** *Evidence before this study:* We conducted two Pubmed searches for reports published after Jan 1, 2000, in all languages. The first search terms were [bacteriophage OR phage] AND [pathogen] AND [diagnostic test]. The primary search criteria identified 57 publications. A secondary criterion excluded reviews and papers on phage display technology. Among twenty remaining articles, bacteriophage were used as proxies for pathogens within diverse genera (Yersinia, Mycobacterium, Burkholderia, Staphylococcus, Salmonella, Bacillus, Escherichia, Acinetobacter, Vibrio); phage profiles were also used as biomarkers for infection. The second search terms were [rapid diagnostic test OR RDT] AND [Vibrio]. After excluding reviews, 101 papers were identified that covered immunologic (antibody-based lateral flow assays), molecular (PCR, qPCR, nl-qPCR) and mass spectrometry-based assays; only 5 articles related to vibriophage detection of which three were permutations of the same initiative.

*Added value of this study:* To our knowledge, this is the first study to investigate the vulnerabilities of RDT performance (e.g., sensitivity and specificity) to virulent bacteriophage and antibiotics in a large multi-site study. The hypothesis that antimicrobials and disease severity might impact RDT performance by decreasing or increasing the target number, respectively, is not novel. However, there is a lack of literature from large clinical studies that rigorously tests this hypothesis. Therefore, the added value of this study is a pragmatic evaluation of this hypothesis and a proof of concept for how to test these complex questions for other less tractable diseases.

*Implications of all the available evidence:* Inside the cholera field, we applied the findings by estimating that changing to a mode of restricted RDT use in which RDTs are reserved for severely dehydrated patients who report no antibiotic exposure, significantly improves the sensitivity. Furthermore, we estimate that an additional 17% of cases would be missed by RDT if the case definition were expanded to include phage detection as a proxy for the pathogen. In this context, the RDT remains with limitations that require consideration when optimizing global RDT deployments. Implications likely extend to other diseases where diagnostics share similar vulnerabilities.

## INTRODUCTION

Cholera causes an estimated 1.3 – 4 million symptomatic cases and 21,000 – 143,000 deaths annually^1^. Since the COVID-19 pandemic, cholera outbreaks expanded to over 44 countries^2^ and the World Health Organization (WHO) declared a global emergency at its highest level (grade 3)^3^. In both endemic and non-endemic regions facing this crisis, access to ‘gold-standard’ culture and molecular based diagnostics at conventional laboratories is limited which can impede effective response. Accurate bedside rapid diagnostic tests (RDTs) offer a promising solution to this problem. In response, international agencies have subsequently operationalized large RDT deployments^4^ for epidemiologic applications that include surveillance and optimization of vaccination campaigns^5,6^.

Cholera is an acute watery diarrhoeal disease caused by the Gram-negative facultative anaerobe *Vibrio cholerae (Vc)*. Case fatality rates historically rose above 20% before intravenous fluids, but in the modern era can fall to less than 1% with use of oral rehydration solution (ORS) alone^7^. The current paradigm is that susceptible individuals become infected after exposure to contaminated food, fomites, or water. The exact infectious dose in humans is unknown and likely varies. In an infant mouse model, the infectious dose may be as low as 20 cells when the inoculum is human-shed *Vc*^8^. The infection may manifest in asymptomatic or symptomatic disease. The ratio of symptomatic to asymptomatic cases is thought to vary widely. In the endemic setting of Bangladesh, it is estimated there are 820 asymptomatic infections (or exposures) per symptomatic case^5^.

Factors that influence cholera severity across the asymptomatic to symptomatic spectrum includes the degree of prior exposure to *Vc*^7^, antimicrobial exposures^9^, and effective predation by virulent bacteriophage (phage)^10^. The WHO Global Task Force on Cholera Control (GTFCC) recommends antibiotic treatment (doxycycline (DOX), azithromycin (AZI), or ciprofloxacin (CIP)) for more severe cases as a supportive maneuver ^11–14^. In practice, mass spectrometry revealed that nearly all cholera patients in a setting like Bangladesh shed diverse antibiotics despite only half reporting exposure^10,15,16^. Antibiotic resistance is common and spreads globally as cholera spreads^17,18^. Virulent phages specific to *Vc* are relatively common and limited in number (ICP1, 2, 3)^19–23^. *Vc* can harbor arrays of phage defense mechanisms, including BREX, on a mobile integrative and conjugative element (ICE) which also harbors antibiotic resistance factors. Phage in turn can harbor cognate antidefenses^24–27^. Phage can also act synergistically or antagonistically with antibiotics to kill bacterial prey^28,29^. While a phenotype called hyper-infectivity is thought to accelerate outbreaks, phage predation is proposed to quench outbreaks although data are limited^30–35^.

Culture and molecular (e.g., PCR, qPCR) assays are considered ‘gold-standards’ for cholera diagnostics^36,37^. Adjunct commercialized RDTs typically are lateral flow devices (‘dipsticks’) that rely on O-specific polysaccharide (OSP) specific antibodies (Abs)^37–39^. Guidelines restrict RDTs to epidemiologic and not clinical applications because of variable performance with sensitivity ranging from 58-100% and specificity from 71- 100%^37,40–42^. Attempts to increase RDT performance by altering protocols and assembly have had limited gains to extend their use to clinical decision making^40,41,43^. Explanations for the varied sensitivity include variations in disease severity, antimicrobial exposure, and phage predation^36,37,9,44^. We previously found that the odds of RDT positivity among cholera patients decreased by more than 80% when phage or antibiotics were detected^9^. This vulnerability calls into question the value of RDTs in their current form and method of use. Determining antimicrobial effects on performance may offer opportunities to optimize deployment and extend use to clinical decision-making.

Our objective herein was to evaluate two primary hypotheses: (i) Virulent phage and antibiotic exposure negatively impact RDT performance. (ii) RDT performance positively correlates with disease severity, defined using the WHO dehydration classifications of mild, moderate and severe^45^. These two hypotheses are connected by target concentration. The mechanisms might be a decrease in *Vc* targets below the RDT limit of detection (LOD) for the first hypothesis versus an increase in *Vc* targets above the LOD for the second hypothesis ^10,46^. To test these hypotheses, we analyzed data from a prospective clinical study conducted across Bangladesh whereby stool samples were collected from patients admitted with diarrhoeal disease at district hospitals^47^. Clinical metadata were paired with microbiologic and molecular data to detect and quantify *V. cholerae* and associated phage; mass spectrometry was conducted on select samples to objectively assess antibiotic effects. The results support our hypotheses exposing critical RDT limitations while also revealing potential approaches that could optimize global RDT deployments.

## MATERIALS AND METHODS

### Ethics Statement

Samples, and associated clinical data, were collected within two previously published IRB approved clinical studies in Bangladesh: (i) The National Cholera Surveillance (NCS) study (icddr,b ERC/RRC PR-15127)^48^ and (ii) The mHealth Diarrhea Management (mHDM) cluster randomized controlled trial (IEDCR IRB/2017/10; icddr,b ERC/RRC PR-17036; University of Florida IRB 201601762) ^47^.

### Study Design

The parent study for this secondary analysis was a prospective longitudinal study of patients admitted with diarrhea at five Bangladesh Ministry of Health and Family Welfare district hospitals (both NCS and mHDM sites) and two centralized NCS hospitals (BITID; icddr,b) (**Fig S1)**. Sites were distributed nation-wide^49^. Samples were collected from March 2018 to December 2018. Samples were collected until April 2019 at one site (icddr,b) where all assays were performed except RDTs.

### Laboratory procedures

Stool samples were collected at admission. Aliquots were stabbed into Cary-Blair transport media for subsequent culture at the icddr,b central laboratory via standard methods^50^. Aliquots for molecular analysis were preserved in 75% RNAlater (Invitrogen); total nucleic acid (tNA) was extracted using standard methods. Criteria and methodology for liquid chromatography-tandem mass spectrometry (LC-MS/MS) to objectively assess antibiotics among all culture positive and select culture negative samples were described previously^10^. In brief, criteria were (i) culture positive for *Vc*, (ii) phage (ICP1,2,3) positive by PCR among culture-negative samples, or (iii) *Vc* positive by PCR (*ompW* target) among a random 10% of those samples negative by culture for *Vc* and by PCR for phage (**Fig S2**). Common antibiotics targeted for LC-MS/MS were based on prior research^10,15^. Ciprofloxacin, doxycycline/tetracycline, and azithromycin, metronidazole and nalidixic acid were analyzed qualitatively. For quantitative analysis of ciprofloxacin, doxycycline/tetracycline, and azithromycin, standard curves were prepared by dilution series of the native and isotopic forms of targets. Each stool sample was spiked with the isotopic mixture as an internal reference control. Quantitative results were binned by effective concentration and ineffective concentration; the definition of effective concentration was based on the lowest minimal inhibitory concentration (MIC) in a prior strain collection^51^. To provide additional insight after the PCR analysis, quantitative PCR (qPCR) was subsequently performed on all available samples to enumerate absolute abundances of *Vc* (*tcpA*), ICP1 (GP58.2), ICP2 (GP24), and ICP3 (GP19) using methods previously described^10^. Estimates for CFU/ml were derived using the qPCR Ct values from standard curves of serial dilutions of cultured *V. cholerae.* Stool samples were tested with the Cholkit (Incepta Pharmaceuticals, Bangladesh), an RDT that targets the detection of *V. cholerae* O1 OPS using a standardized method^39,52^. In brief, five drops of watery stool were added to the diluent supplied in a microcentrifuge tube. The RDT strip was then dipped into the stool-diluent mixture for 15 minutes. *V. cholerae* O1 detection was confirmed ‘by eye’ for the appearance of both test and control lines^39^. Solid stools were not tested; cholera RDTs in general, including the Cholkit, are not designed to test solid stools.

### Statistical analyses

Statistics and visualizations were done in GraphPad Prism version 10.2.3 and R version 4.4.2^53^. The “Eulerr” package was used to create **Figure 1A** for samples with all relevant test results available^54^. Test performance metrics (sensitivity, specificity, positive predictive value, negative predictive value) and corresponding exact binomial confidence limits were calculated in the “epiR” package^55^. Standardized predictive values were calculated by using the observed diagnostic sensitivity and specificity in the given subgroup and calculating the predictive value that would have been observed if disease prevalence was 50% in that subgroup. We used unadjusted generalized linear models with a binomial error distribution and logit link to estimate odds ratios and corresponding 95% confidence intervals. Fisher’s exact test was used to calculate p-values where indicated.

**Fig. 1.**
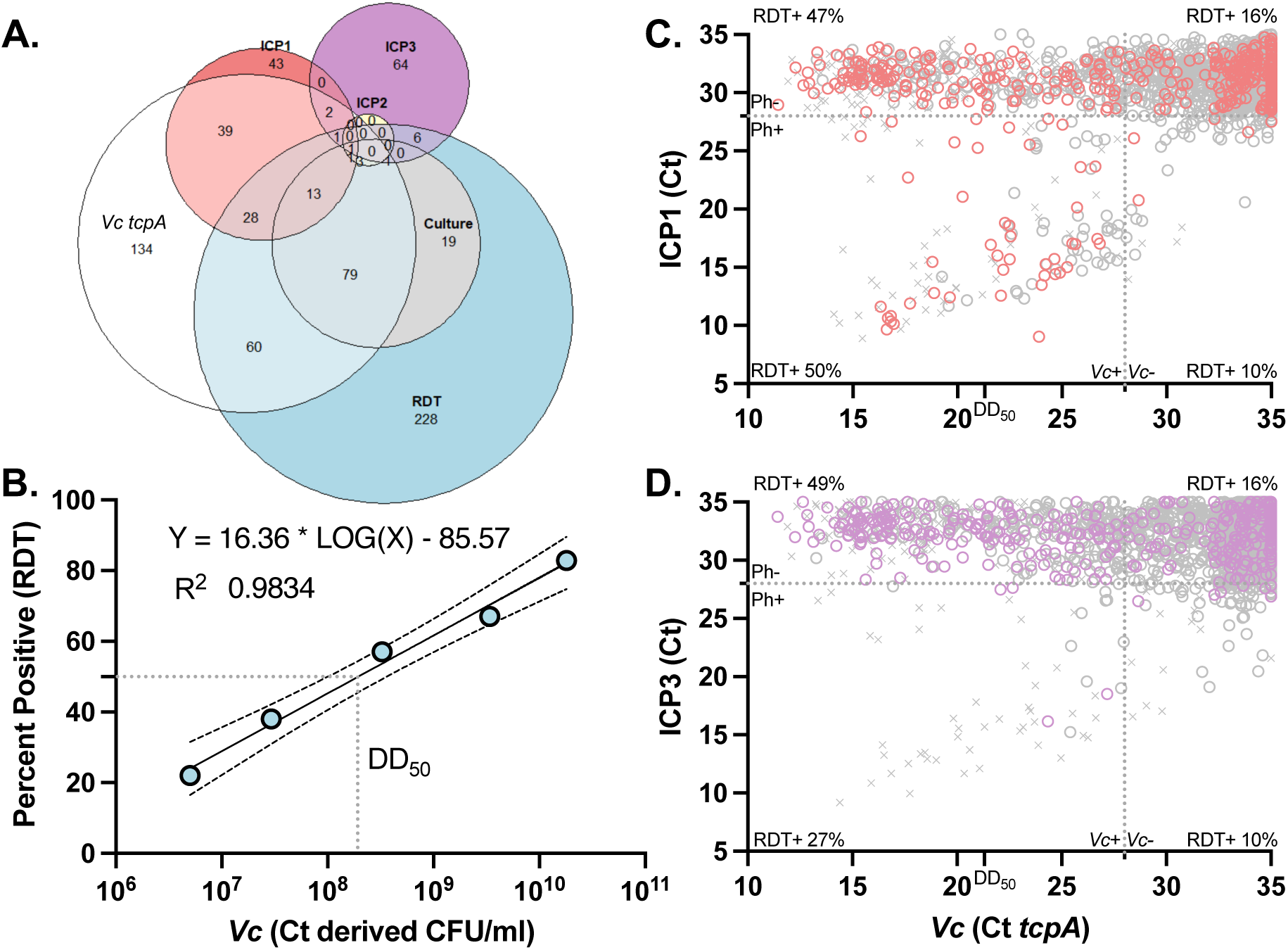
Comparison of target detection by different diagnostics. **A** Euler plot displaying overlaps between positive diagnostics. Given limitations with Euler plots, the following positive values are not shown in the figure: only Vc culture (n= 35/175 total culture positives; 25%), only ICP2 qPCR (n=4/10 total ICP2 positives; 40%), only Vc culture and Vc qPCR tcpA (n=18), only RDT and ICP1 qPCR (n=4), and only Vc qPCR tcpA and ICP3 qPCR (n=9). **B**. Linear regression analysis of relationship between log_10_ *Vc* abundance (CFU/ml) derived from *tcpA* qPCR and RDT positivity. Samples were binned into five groups based on *Vc* concentration: less than 1x10^7^, 1-9 x 10^7^, 1-9 x 10^8^, 1-9 x 10^9^, and above 1x10^10^ CFU/ml. In a given group, each symbol is plotted by the median concentration (X coordinate) by the proportion of samples RDT positive (Y coordinate). The *Vc* dose at which 50% of RDTs (‘DD_50_’) were positive (gray dotted line at 1.93 x10^8^ CFU/ml). Line of best fit is shown and fit using least squares regression (95% CIs shown). **C.** Comparison of qPCR Ct values for *Vc* (*tcpA*) with ICP1. **D.** Comparison of qPCR Ct values for *Vc* (*tcpA*) with ICP3 (GP19). For C and D: Quadrants are demarcated by phage (Ph) and *Vc* positivity (+) with the percentage RDT positive per quadrant. Gray dotted lines = thresholds for positive (Ct = 28). “X” = no RDT performed; hollow colored circle = RDT+, hollow gray circle = RDT-. Tick shows dose at which 50% of all RDTs are positive (DD_50_).

## RESULTS

### Study overview

A total of 2574 participants were enrolled at seven hospitals across Bangladesh from March to December 2018, with one stool sample collected from each participant (**Figs S1 and S2**). Enrollment continued at one site (icddr,b) until April 2019. Although RDTs were not performed at the icddr,b, samples were included to better assess phage and *Vc* relationships by qPCR. RDTs were performed on each of the remaining 2180 samples of which all had culture and 2078 had *Vc* qPCR **(Table 1)**. Specific to testing our two hypotheses: (i) ICP1, ICP2, and ICP3 were detected among 137, 10, and 86 participants, respectively, and (ii) the number of participants with mild, moderate and severe dehydration was 392, 1193 and 493, respectively. LC-MS/MS for antibiotic detection was attempted and successful on all culture positive samples and a subset of culture negative samples (446) ^10,15,51^ **(Fig S2).**

**Table 1.**
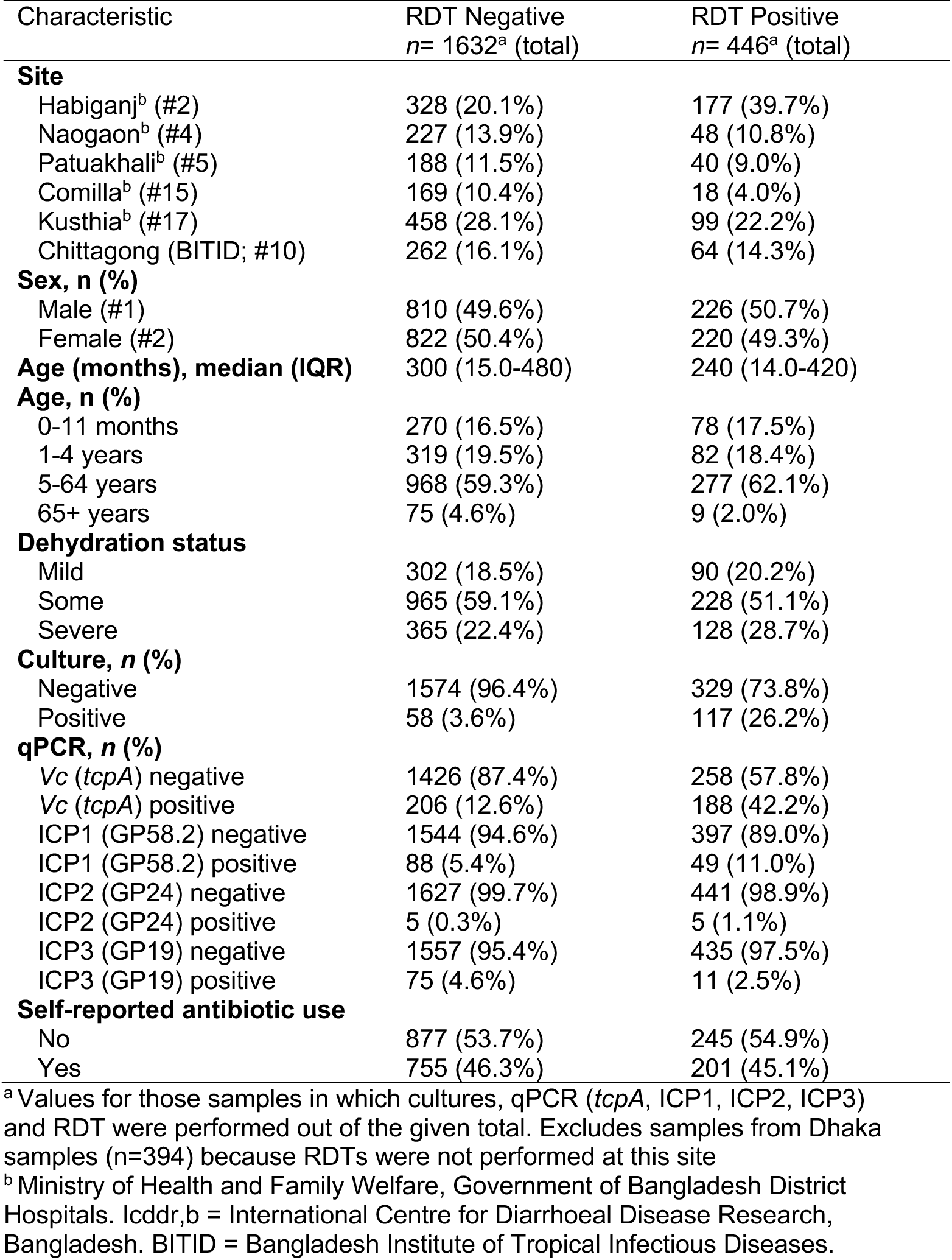
Overview of participants characteristics by RDT result.

### Characterization and comparison of assays for *Vc* detection

Among samples with all assays performed, there was discordance in *Vc* detection by assay type (**Fig 1A**). Out of 2078 samples with both *Vc* culture and qPCR, only 5.8% (120) of samples were positive for *Vc* by both culture and qPCR. An additional 13.2% (274) of samples were negative by culture and positive by qPCR, and 2.6% (55) of samples were positive by culture and negative by qPCR. The concentration of *Vc* at which 50% of the RDTs tested positive (diagnostic dose 50 or ‘DD_50_’) was 1.93 x 10^8^ CFU/ml; The percentage of positive RDTs increased with *Vc* concentration binned by order of magnitude (**Fig 1B**).

### Distribution of phage among samples with and without *Vc*

Among samples with all assays performed, ICP1, ICP2 and ICP3 were detected by qPCR in 137, 10 and 76 samples, respectively (**Table 1**). These analyses include samples that were positive or negative for *Vc*. In the absence of *Vc* (culture or qPCR), ICP1, ICP2, and ICP3 were detected in 2.1% (43), 0.2% (4), and 3.1% (64) of samples respectively (**Fig 1A**); unless specified, analyses focused on ICP1 and ICP3 because of the larger sample sizes. We next analyzed quantitative associations between phage and *Vc.* Data are visualized in quadrants by *Vc* and/or phage detection using a Ct threshold of 28 (**Fig 1C, D**). Samples that did not have RDT results (gray ‘x’) were included to better elicit patterns in the data. For both ICP1 and ICP3, increasing *Vc* detection was associated with increasing phage detection. Among those samples with RDT results, the percentages of RDT positive samples are shown in each quadrant. The quadrants with the least percentage of RDT positive samples were those with phage detection alone.

### Effect of phage predation on RDT performance

Among cholera samples defined conventionally by *Vc* qPCR or culture, RDT sensitivity (46%) and specificity (85%) did not differ when phage positive samples were excluded (**Table 2**). PPV and NPV were respectively higher and lower among samples with ICP1 compared to other groups (e.g., samples without phage). However, this effect was driven by differences in cholera prevalence between groups, and resolved once predictive values were assessed at a standardized prevalence. Among samples with ICP3, sensitivity decreased compared to samples without phage yet confidence intervals overlapped. While culture and qPCR are the current reference standards, the detection of cholera-specific phage among samples negative for *Vc* suggested the conventional definition may inappropriately exclude cases. Therefore, we explored an expanded definition that included phage detection as a proxy for *Vc* for situations when the primary *Vc* target might be highly degraded. The inclusion of phage in the diagnostic criteria resulted in a modest effect on the RDT (**Table S1**); the sensitivity decreased from 0.46 (95%CI 0.41–0.51) to 0.38 (95%CI 0.34-0.42). Despite this seemingly small effect on sensitivity, we estimated RDTs would miss an additional 17% of ‘true’ cholera cases if the diagnostic reference standard were expanded to include “phage only” as cholera cases.

**Table 2.** Effect of phage predation on RDT performance.

| Sample type | All | No phage<sup>a</sup> | ICP1 phage positive | ICP3 phage positive |
| --- | --- | --- | --- | --- |
| Vc+/total (%)<sup>b</sup> | 449 / 2078 (21.6) | 345 / 1852 (18.6) | 89 / 137 (65.0) | 15 / 86 (17.4) |
| Sensitivity | 0.46 (0.41, 0.51) | 0.46 (0.40, 0.51) | 0.49 (0.39, 0.60) | 0.27 (0.08, 0.55) |
| Specificity | 0.85 (0.84, 0.87) | 0.85 (0.83, 0.87) | 0.90 (0.77, 0.97) | 0.90 (0.81, 0.96) |
| PPV<sub>obs</sub><sup>c</sup> | 0.46 (0.42, 0.51) | 0.41 (0.36, 0.46) | **0.90 (0.78, 0.97)** | 0.36 (0.11, 0.69) |
| NPV<sub>obs</sub><sup>c</sup> | 0.85 (0.83, 0.87) | 0.87 (0.85, 0.89) | **0.49 (0.38, 0.60)** | 0.85 (0.75, 0.92) |
| PPV<sub>50</sub><sup>c</sup> | 0.76 (0.72, 0.79) | 0.75 (0.71, 0.79) | 0.83 (0.68, 0.93) | 0.73 (0.45, 0.92) |
| NPV<sub>50</sub><sup>c</sup> | 0.61 (0.59, 0.64) | 0.61 (0.58, 0.64) | 0.64 (0.53, 0.73) | 0.55 (0.43, 0.67) |

<sup>a</sup> ICP1, ICP2, and ICP3 not detected
<sup>b</sup> Number positive for Vc by culture or qPCR (*tcpA*) as the gold reference standard over total number samples in grouping
<sup>c</sup> Subscript indicates the proportion of samples that were gold-standard positive in the given group for the calculation of each predictive value, with "obs"=observed and "50"=50%. For example, PPV<sub>50</sub> or standardized positive predictive value indicates the positive predictive value of RDT in each group if 50% of that group was gold-standard positive for cholera.

### Effect of dehydration severity on RDT performance

We evaluated the impact of dehydration severity on RDT performance. The median concentrations of *Vc* (CFU/ml) derived from those samples positive by qPCR at mild, moderate or severe levels of dehydration were 3.25 x 10^7^ (IQR 8.60 x 10^6^ to 1.14 x 10^9^), 1.05 x 10^8^ (IQR 1.06 x 10^7^ to 2.16 x 10^9^) and 1.00 x 10^8^ (IQR 8.34 x 10^6^ to 4.41 x 10^9^), respectively. In this context, RDT performance increased sequentially from patients with mild to moderate to severe dehydration (**Table 3**). RDT sensitivity was 0.25 (95% CI: 0.16, 0.37) in those mildly dehydrated, compared to 0.63 (95% CI: 0.54, 0.71) in samples from severely dehydrated patients. Specificity also increased between mild cases and cases with moderate or severe dehydration. Standardized predictive values, both positive and negative, also increased significantly in severely dehydrated patients compared to mildly dehydrated patients.

**Table 3.** Effect of dehydration severity on RDT performance.

|  | Mild (comparator)<sup>a</sup> | Moderate<sup>a</sup> | Severe<sup>a</sup> |
| --- | --- | --- | --- |
| Vc+/total (%)<sup>b</sup> | 75 / 392 (19.1)<sup>c</sup> | 236 / 1193 (19.8)<sup>c</sup> | 138 / 493 (28.0)<sup>c</sup> |
| Sensitivity | 0.25 (0.16, 0.37) | 0.43 (0.36, 0.49) | **0.63 (0.54, 0.71)** |
| Specificity | 0.78 (0.73, 0.82) | **0.87 (0.84, 0.89)** | **0.88 (0.85, 0.92)** |
| PPV<sub>obs</sub><sup>d</sup> | 0.21 (0.13, 0.31) | **0.44 (0.38, 0.51)** | **0.68 (0.59, 0.76)** |
| NPV<sub>obs</sub><sup>d</sup> | 0.81 (0.77, 0.86) | 0.86 (0.84, 0.88) | 0.86 (0.82, 0.89) |
| PPV<sub>50</sub><sup>d</sup> | 0.53 (0.42, 0.63) | **0.76 (0.71, 0.81)** | **0.85 (0.78, 0.89)** |
| NPV<sub>50</sub><sup>d</sup> | 0.51 (0.45, 0.57) | **0.60 (0.57, 0.64)** | **0.71 (0.65, 0.76)** |

<sup>a</sup> Mild, moderate and severe dehydration approximate 0-4%, 5-9% and 10%+ loss to normal body due to diarrhoeal fluid loss per WHO guidelines.
<sup>b</sup> Number positive for Vc by culture or qPCR (*tcpA*) as the gold reference standard over total number samples in grouping
<sup>c</sup> The concentration of Vc derived from qPCR data among patients with mild, moderate and severe dehydration is $3.25 \times 10^7$ (IQR $8.60 \times 10^6$ to $1.14 \times 10^9$ ), $1.05 \times 10^8$ ( $1.06 \times 10^7$ to $2.16 \times 10^9$ ) and $1.00 \times 10^8$ ( $8.34 \times 10^6$ to $4.41 \times 10^9$ ), respectively.
<sup>d</sup> Defined in Table 2.

### Effect of phage on the RDT performance at different dehydration levels

We explored if the effect of phage on RDT performance varied by dehydration status (possible effect modification) **(Table 4)**. Among *Vc* positive patients (culture or qPCR), phage exposure was not associated with RDT positivity as an aggregate or by severity level. When the case definition was expanded to include phage detection, the odds of RDT positivity were significantly lower in those with phage compared to those without phage (OR 0.377; 95%CI 0.217-0.642). This effect was observed among samples from moderately and severely dehydrated cases, but not from mildly dehydrated cases. The effect also differed by phage type used in the case definition. The effect of phage on RDT positivity was most prominent among those samples positive by culture, qPCR, or ICP3 (OR 0.099; 95%CI 0.023-0.292). The impact of dehydration status on this relationship between phage presence and RDT performance (effect modification) was also strongest with this expanded definition.

**Table 4.** Effect of phage exposure on RDT positivity among cholera patients.

| Table 1. Effect of phage exposure on RDT positivity among cholera patients |  |  |  |  |  |  |
| --- | --- | --- | --- | --- | --- | --- |
| Case definition | n | % RDT positive (n/ total) |  | OR<sup>a</sup> | CI<sup>a</sup> | P value<sup>a</sup> |
|  |  | Phage Exposed | Unexposed |  |  |  |
| Vc positive<sup>a</sup> |  |  |  |  |  |  |
| All | 197 | 46 (22/48) | 46 (69/149) | 0.981 | 0.508-1.88 | 0.954 |
| Mild | 43 | 6 (1/16) | 15 (4/27) | 0.383 | 0.019-2.91 | 0.411 |
| Moderate | 86 | 57 (8/14) | 42 (30/72) | 1.867 | 0.589-6.21 | 0.291 |
| Severe | 68 | 72 (13/18) | 70 (35/50) | 1.114 | 0.349-3.97 | 0.859 |
| Vc positive or phage positive (ICP1,2,3)<sup>a</sup> |  |  |  |  |  |  |
| All | 259 | 25 (27/110) | 46 (69/149) | **0.377** | **0.217-0.642** | **<0.001** |
| Mild | 62 | 14 (5/35) | 15 (4/27) | 0.958 | 0.229-4.25 | 0.953 |
| Moderate | 113 | 20 (8/41) | 42 (30/72) | **0.339** | **0.130-0.809** | **0.019** |
| Severe | 84 | 41 (14/34) | 70 (35/50) | **0.300** | **0.118-0.737** | **0.010** |
| Vc positive or ICP1 positive<sup>a</sup> |  |  |  |  |  |  |
| All | 218 | 32 (22/69) | 46 (69/149) | **0.543** | **0.294-0.980** | **0.046** |
| Mild | 57 | 13 (4/30) | 15 (4/27) | 0.885 | 0.189-4.13 | 0.872 |
| Moderate | 93 | 33 (7/21) | 42 (30/72) | 0.700 | 0.240-1.90 | 0.494 |
| Severe | 68 | 61 (11/18) | 70 (35/50) | 0.673 | 0.220-2.14 | 0.491 |
| Vc positive or ICP3 positive<sup>a</sup> |  |  |  |  |  |  |
| All | 187 | 8 (3/38) | 46 (69/149) | **0.099** | **0.023-0.292** | **<0.001** |
| Mild | 32 | 20 (1/5) | 15 (4/27) | 1.44 | 0.065-13.6 | 0.770 |
| Moderate | 91 | 5 (1/19) | 42 (30/72) | **0.078** | **0.004-0.409** | **0.015** |
| Severe | 64 | 7 (1/14) | 70 (35/50) | **0.033** | **0.002-0.188** | **0.002** |

<sup>a</sup>Reference standard to define positive cholera cases for analysis within a given group. "Vc positive" is defined as positive by culture or qPCR. *P* values were computed using logistic regression (see methods).

### Effect of antibiotic exposure on RDT performance

Large discordance between self-reported and LC-MS/MS-detected antibiotic exposures was observed. Among those with complete diagnostics, 46% of patients (956/2078) self-reported antibiotic exposure and 87% (220/252) of select samples assayed by LC-MS/MS (**Fig S2**) contained antibiotics. Among the WHO recommended antibiotics, ciprofloxacin was the most commonly self-reported antibiotic (27%; 568) followed by azithromycin (14%; 289) with no participant self-reporting doxycycline use (**Table S2**). RDT performance was consistent across all types of self-reported antibiotic exposure, including no self-reported antibiotic exposure. To take a more objective approach, LC-MS/MS was used to detect exposure to common antibiotics (CIP, AZI, DOX, metronidazole (MET), or nalidixic acid (NAL) which were detected in 81% (178/220) of samples tested (**Table S2**). The small proportion of samples that did not have antibiotics detected by LC-MS/MS limited analyses on the effects of antibiotics on RDT performance. Despite this challenge, RDT sensitivity among samples with AZI at or above the ‘effective concentration’ was 0.33 (95%CI 0.13-0.59) compared to 0.67 (95%CI 0.47-0.83) among samples without antibiotic (**Table S2**). Among *Vc* positive samples, antibiotic resistance had no effect on the odds of RDT positivity with or without AZI exposure, however samples sizes were small (**Table S3**).

### Estimates of RDT performance in different modes of deployment

We sought to estimate and compare RDT performance in different modes of deployment (**Table 5**). We considered an unrestricted mode in which all patients are tested. We compared this to a more restricted mode that would rely on data accessible at the bedside through the clinical history (self-reported antibiotic exposure) or the physical exam (dehydration status). Despite phage detection not being accessible at the bedside, it is reasonable to consider it as a potential component to include in molecular diagnostic panels in the future. LC-MS/MS is too expensive and complex for consideration. In this context, we found that RDT sensitivity increases from 0.46 (95%CI: 0.41-0.51) among all patients to 0.69 (95%CI: 0.57-0.80) when use is restricted to patients that report no antibiotic exposure and have severe dehydration. When the case definition is expanded to include phage, RDT sensitivity also increases between restricted and unrestricted modes. Modest effects were observed on PPV and NPV depending on case definition and mode of use.

**Table 5.** Estimates of RDT performance in different modes of deployment.

| Approach | Unrestricted<br>(comparator) | Use clinical<br>exam | Use patient history<br>and exam | Use clinical<br>exam | Use patient history<br>and exam |
| --- | --- | --- | --- | --- | --- |
| Restriction | None | Moderate and<br>severe cases | Moderate and<br>severe cases AND<br>no reported<br>antibiotic use | Severe cases | Severe cases AND<br>no reported<br>antibiotics use |
| Case definition (current gold standard): culture or qPCR for Vc |  |  |  |  |  |
| Cases/total (%) | 449 / 2078 (21.6) | 374 / 1686 (22.2) | 168 / 863 (19.5) | 138 / 493 (28.0) | 72 / 266 (27.1) |
| Sensitivity | 0.46 (0.41, 0.51) | 0.50 (0.45, 0.55) | 0.54 (0.46, 0.61) | **0.63 (0.54, 0.71)** | **0.69 (0.57, 0.80)** |
| Specificity | 0.85 (0.84, 0.87) | 0.87 (0.85, 0.89) | 0.86 (0.83, 0.89) | 0.88 (0.85, 0.92) | 0.86 (0.80, 0.91) |
| PPV<sub>obs</sub><sup>b</sup> | 0.46 (0.42, 0.51) | 0.53 (0.47, 0.58) | 0.48 (0.41, 0.56) | **0.68 (0.59, 0.76)** | **0.65 (0.53, 0.75)** |
| NPV<sub>obs</sub><sup>b</sup> | 0.85 (0.83, 0.87) | 0.86 (0.84, 0.88) | 0.88 (0.86, 0.91) | 0.86 (0.82, 0.89) | 0.88 (0.83, 0.93) |
| PPV<sub>50</sub><sup>b</sup> | 0.76 (0.72, 0.79) | 0.80 (0.76, 0.83) | 0.79 (0.74, 0.84) | 0.85 (0.78, 0.89) | 0.83 (0.75, 0.90) |
| NPV<sub>50</sub><sup>b</sup> | 0.61 (0.59, 0.64) | 0.64 (0.61, 0.66) | 0.65 (0.61, 0.69) | **0.71 (0.65, 0.76)** | **0.74 (0.66, 0.81)** |
| Case definition (expanded standard): culture or qPCR for Vc OR qPCR for ICP1/ICP3 phage |  |  |  |  |  |
| Cases/ total (%) | 567 / 2078 (27.3) | 469 / 1686 (27.8) | 216 / 863 (25.0) | 173 / 493 (35.1) | 89 / 266 (33.5) |
| Sensitivity | 0.38 (0.34, 0.43) | 0.41 (0.37, 0.46) | 0.43 (0.36, 0.50) | **0.51 (0.43, 0.59)** | **0.57 (0.46, 0.68)** |
| Specificity | 0.85 (0.83, 0.87) | 0.87 (0.85, 0.89) | 0.85 (0.82, 0.88) | 0.88 (0.83, 0.91) | 0.85 (0.79, 0.90) |
| PPV<sub>obs</sub><sup>b</sup> | 0.49 (0.44, 0.54) | 0.54 (0.49, 0.60) | 0.49 (0.42, 0.57) | **0.69 (0.60, 0.77)** | **0.66 (0.55, 0.77)** |
| NPV<sub>obs</sub><sup>d/b</sup> | 0.79 (0.77, 0.81) | 0.79 (0.77, 0.81) | 0.82 (0.79, 0.85) | 0.77 (0.72, 0.81) | 0.80 (0.73, 0.85) |
| PPV<sub>50</sub><sup>b</sup> | 0.72 (0.68, 0.76) | 0.76 (0.71, 0.80) | 0.74 (0.68, 0.80) | 0.80 (0.73, 0.86) | 0.80 (0.70, 0.87) |
| NPV<sub>50</sub><sup>b</sup> | 0.58 (0.55, 0.60) | 0.60 (0.57, 0.62) | 0.60 (0.56, 0.64) | 0.64 (0.59, 0.69) | 0.67 (0.59, 0.74) |

<sup>a</sup> Shaded text represents the current unrestricted standard of RDT use using qPCR and culture for Vc as the reference standard. Bold text compares performance horizontally between unrestricted and restricted approaches; underlined text vertically compares sensitivity when the reference standard is expanded to include the detection of phages by qPCR (ICP1/3)
<sup>b</sup> Defined in Table 2.

## DISCUSSION

Effective cholera control is aided by accurate bedside rapid diagnostic tests (RDTs). Here we conducted a secondary analysis of a prospective cohort study of patients with diarrhoeal disease to test if virulent bacteriophage (phage) impact RDT performance and determine if there are tractable approaches to optimize RDT use. With the current gold standard of culture or qPCR *Vc* positivity, we found no significant effect of phage on RDT performance. When the reference standard was expanded to include phage, there was a small decrease in RDT sensitivity. In contrast, significantly higher sensitivity and specificity were found among patients with moderate or severe dehydration. Using the expanded definition of including phage detection, the odds of RDT positivity among cholera patients decreased by 57% with phage exposure and the effect was most robust among patients with severe dehydration. These findings align with our guiding hypotheses as well as pose intriguing questions on how one might apply these results to optimize RDT deployments.

Our first hypothesis was that virulent phage and antibiotic exposure negatively impact RDT diagnostic performance. RDT sensitivity and specificity were not appreciatively impacted when samples with phage were removed from the analysis. Among ICP1 positive samples, PPV and NPV increased and decreased respectively using the current gold standard case definition. This likely reflects an increased pretest probability for *Vc*; standardizing rates of *Vc* removed the effect. Among ICP3 positive samples, sensitivity and PPV were decreased however confidence intervals overlapped; standardizing rates of *Vc* removed the effect on PPV. We next expanded the case definition to include phage detection. Inclusion of phage in the diagnostic criteria had modest effects that aligned with our guiding hypotheses.

Our second hypothesis was that RDT performance positively correlates with disease severity. The rationale is based on our prior study of the predator (phage) -- prey (*Vc*) dynamics along the disease severity spectrum; the prior study found high abundance of *Vc* was correlated with severe disease^10^. In contrast to the modest effects of phage, significant increases in sensitivity and specificity were found among patients with moderate or severe dehydration. This finding likely reflects that samples from more severe patients had higher concentrations of *Vc* at levels that we showed above have a higher percentage of RDT positivity.

The lack of congruence between the assays performed (culture, qPCR, RDTs) may reflect complex underlying biologic events and not simply variation in assay assembly and manipulation. Mechanistically, we propose that false positive RDTs may be more common among mild cases because the concentration of *Vc* is low and non-*Vc* microbiota is high which may increase cross-reactivity of the immunologic epitopes. Although cross-reactivity may occur using molecular assays, cross-reactivity for the qPCR primer set used was likely low based on our prior study^10^; that said degradation of the nucleic acid impacts molecular assays which we attempted to mitigate by stabilizing the samples. Culture is a definitive assay for *Vc* but sensitivity can decrease because of processing delays, antibiotic exposure and possibly phage predation. In this context, we evaluated the interactions between phage predation and disease severity. Using the expanded case definition that includes phage detection, we found that the odds of RDT positivity among cholera patients decreased by 63% with phage exposure. The effect was most robust among patients with severe dehydration and varied by phage type; ICP3 dramatically decreased the odds of RDT positivity among cholera patients. One explanation for this effect of among severely dehydration patients with ICP3 is that there is simply more prey for the predator among severely dehydration patients and the effect is more easily detected. Among patients with mild disease, there may simply be insufficient *Vc* for the RDT to detect independent of phage predation.

Both clinicians and epidemiologists face two key questions based on these findings. The first question is whether to include phage detection in cholera diagnostic panels as a proxy for detection of the pathogen when phage degrade the pathogen below the limit of detection for a given assay. The benefits (e.g. overall increased case detection) versus risks (e.g. increased cost) need consideration by both clinicians and epidemiologists. The second question is how to best validate and incorporate these findings as RDTs are being deployed globally to combat cholera. For validation, studies in diverse clinical and epidemiologic scenarios are needed. For deployment, we estimated the effects of restricting RDT use to patients who report no antibiotic use and have severe dehydration. In this context, sensitivity had a relative increase of approximately 50%, independent of consideration of phage predation. This restriction may represent a strategy to conserve RDTs in resource limited situations. However, this restriction may also miss non-severe disease that play an important role in transmission or mortality in the convalescent period.

These findings should be considered within the limitations of the study. First, LC-MS/MS was not performed on all samples because of logistical limitations. Among *Vc* or phage positive samples, LMCS found antibiotics were common and the distributions of samples were insufficient to disaggregate the effects of phage and antibiotics on RDT performance; proposing to restrict RDT use to those without antibiotic exposure is in some aspects empiric. Second, the recruitment and enrollment of patients within the parent clinical trial was based on the census of patients that sought care. This may a potential recruitment bias. Those patients that sought care are not considered representative of all infected individuals in the community. Third, clinical exams were conducted in district hospitals using WHO and Bangladesh Ministry of Health and Family Welfare guidelines. These guidelines have varied accuracy; this study would likely be improved by incorporating more accurate measures of dehydration^56^. Fourth, the approach had experimental limitations: The analyses excluded host factors such as innate and acquired immunity. Control assays were not performed on asymptomatic/healthy participants in the community. Solid stools were not assayed. Despite these limitations, the findings provide important actionable insights. Future time-series studies of patients over the disease course and recovery period in endemic and non-endemic settings would assist testing our guiding hypotheses and address study limitations.

## CONCLUSION

Herein, we found RDT performance was minimally impacted by phage overall and significantly increased among severely dehydrated patients. Expanding the case definition to include phage, the odds of RDT test positivity decreased significantly. We favor the expanded case definition that includes phage while recognizing the need for future investigation. Lastly, we provide estimates on the applied effects of restricted vs non-restricted RDT deployment. The significance of these findings may extend broadly to other disease where diagnostics share similar vulnerabilities.

## Data availability

Available upon request.

## Code availability

Available upon request.

## Acknowledgements

We thank the patients for their participation. We thank the diligent clinical and laboratory team members who were essential to the success of the study. We are grateful to the Institute of Epidemiology, Disease Control and Research (IEDCR), Ministry of Health and Family Welfare, and Government of Bangladesh who collaborated on the original parent clinical studies. We are also grateful to the Governments of Bangladesh and Canada for providing unrestricted support to the icddr,b. We are grateful to B. Johnson, T. Linn, N. Rushing and K. Berquist for their administrative guidance at the University of Florida.

## Author contributions

Conceptualization: SMA, MAS, FQ, JA, AIK, EJN

Implementation of clinical study: MAS, FQ, AIK, EJN

Lab procedures: MAS, ETC, ACM, KI, MIUK, MTRB, YB, EF, AV, LB, MK, LSB, EJN

Statistical analyses and visualizations: SMA, NM, BJS, JA, EJN

Funding acquisition: SMA, DS, KBS, EJN

Supervision: KBS, BJS, FQ, AIK, EJN

Writing – original draft: SMA, EJN

Writing – review & editing: SMA, MAS, DS, AKD, JA, FQ, AIK, EJN

---

**Figure S1.**
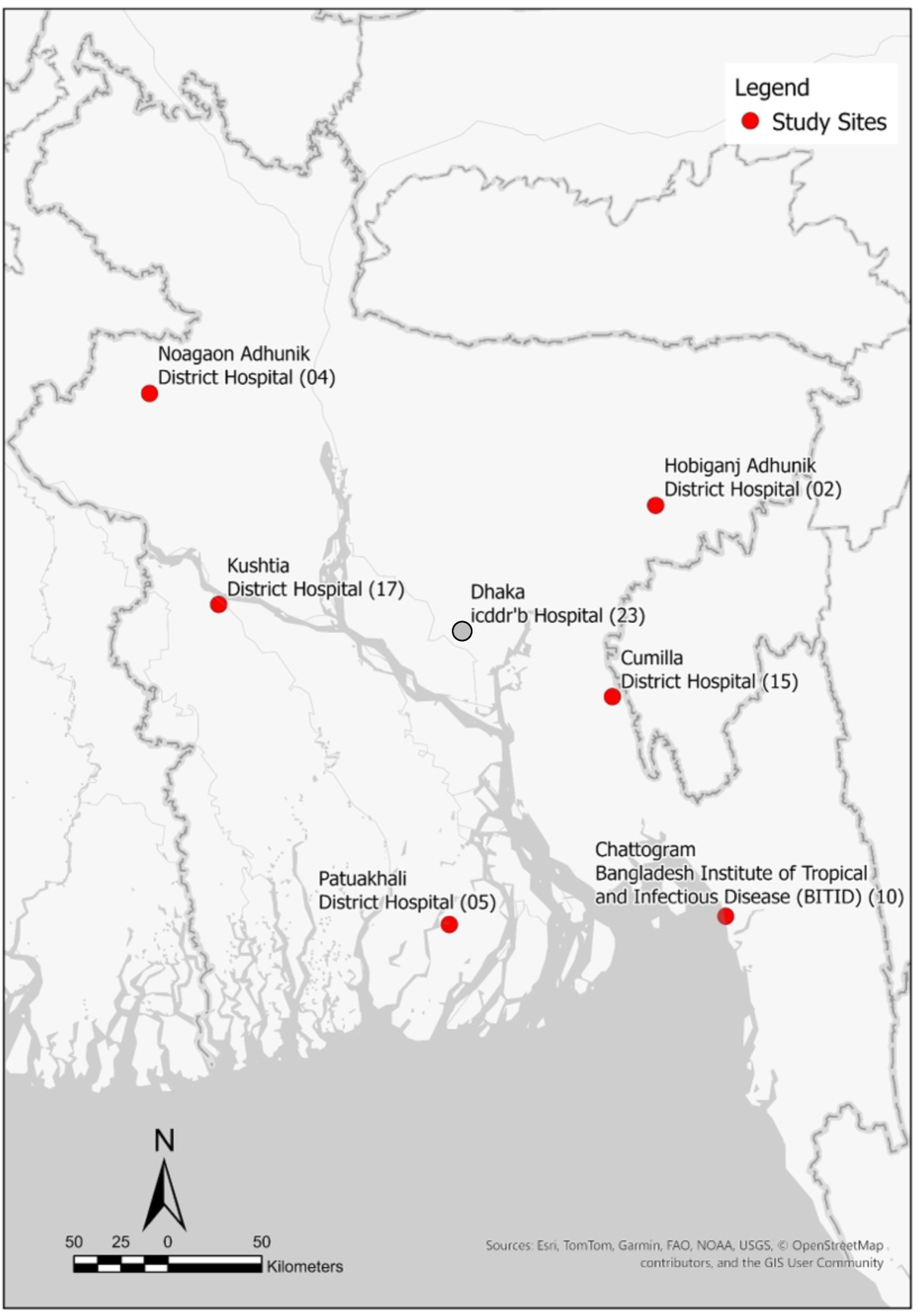
Study Map. Study site locations across Bangladesh. Red circles = RDT performed; gray circle = RDT not performed. Numbers in brackets correlate with site numbers in Table 1.

**Figure S2.**
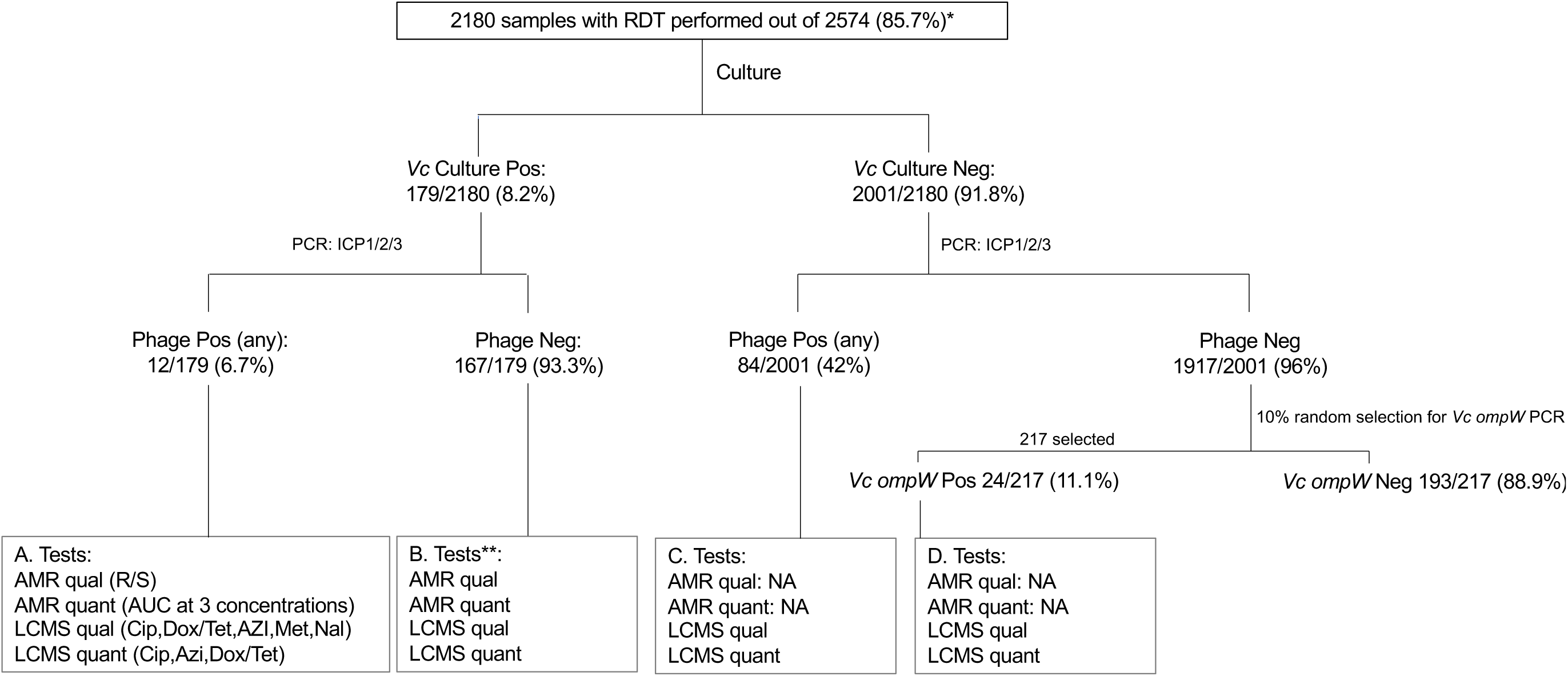
Sample processing diagram. Schematic of the number of samples collected from patients admitted with diarrhoeal disease (one per patient). * Those samples with RDTs (2180) are represented in the flow diagram; RDTs were not performed at the icddr,b. ** Seven samples were excluded from further analysis (AMR testing and LC-MS/MS) because they were misclassified as culture negative.

**Table S1.**
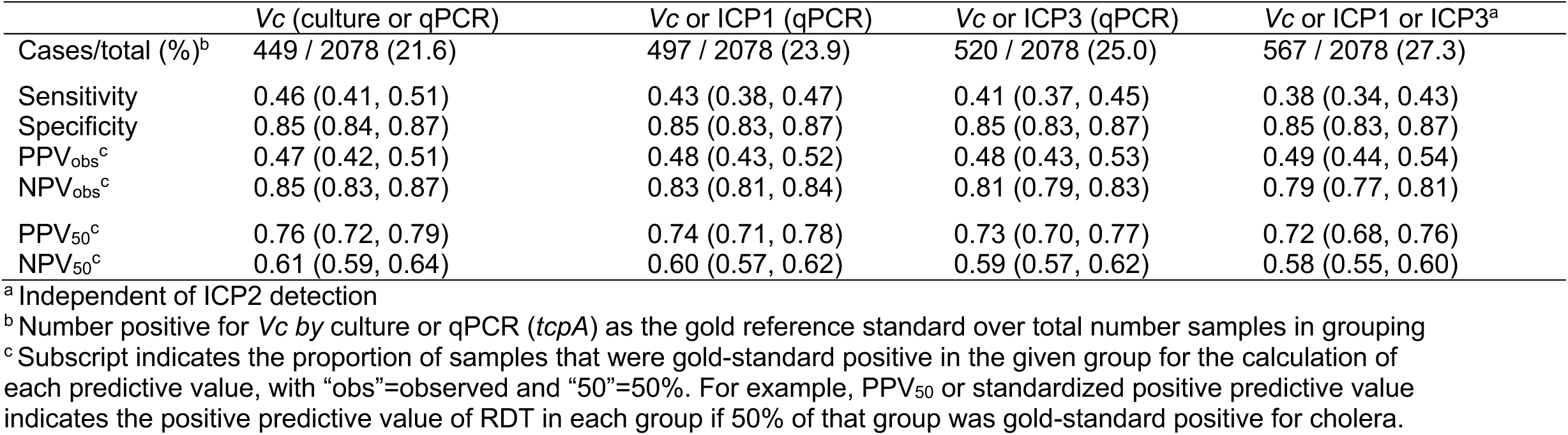
Comparison of RDT performance using different diagnostic criteria for cholera.

**Table S2.** RDT performance as a function of antibiotic exposure.

| Exposure: | Reference<br>No antibiotic | Antibiotic (any)<br>Exposure | Ciprofloxacin |  | Azithromycin |  | Doxycycline/Tetracycline<sup>a</sup> |  |
| --- | --- | --- | --- | --- | --- | --- | --- | --- |
|  |  |  | Reported vs<br>Detected | At effective<br>Concentration<sup>b</sup> | Reported vs<br>Detected | At effective<br>Concentration<sup>b</sup> | Reported vs<br>Detected | At effective<br>Concent.<sup>b</sup> |
| **Self-reported (all patients)** |  |  |  |  |  |  |  |  |
| Vc+/total(%)<sup>c</sup> | 210 / 1122 (18.7) | 239 / 956 (25.0) | 159 / 568 (28.0) | --- | 58 / 289 (20.1) | --- | --- | --- |
| Sensitivity | 0.46 (0.39, 0.53) | 0.46 (0.40, 0.53) | 0.48 (0.40, 0.56) | --- | 0.43 (0.30, 0.57) | --- | --- | --- |
| Specificity | 0.84 (0.81, 0.86) | 0.87 (0.85, 0.90) | 0.89 (0.86, 0.92) | --- | 0.87 (0.82, 0.91) | --- | --- | --- |
| PPV<sub>obs</sub><sup>d</sup> | 0.39 (0.33, 0.46) | 0.55 (0.48, 0.62) | 0.63 (0.54, 0.71) | --- | 0.45 (0.32, 0.59) | --- | --- | --- |
| NPV<sub>obs</sub><sup>d</sup> | 0.87 (0.85, 0.89) | 0.83 (0.80, 0.86) | 0.81 (0.78, 0.85) | --- | 0.86 (0.81, 0.90) | --- | --- | --- |
| PPV<sub>50</sub><sup>d</sup> | 0.74 (0.69, 0.78) | 0.79 (0.73, 0.83) | 0.81 (0.75, 0.87) |  | 0.77 (0.66, 0.85) |  |  |  |
| NPV<sub>50</sub><sup>d</sup> | 0.61 (0.57, 0.64) | 0.62 (0.58, 0.66) | 0.63 (0.58, 0.68) |  | 0.60 (0.53, 0.67) |  |  |  |
| **Detected by Mass Spectrometry (cholera patients only<sup>e</sup>)** |  |  |  |  |  |  |  |  |
| Vc+/total(%)<sup>a</sup> | 30 / 32 (93.8) | 178 / 220 (80.9) | 118 / 148 (79.7) | 86 / 116 (74.1) | 47 / 68 (69.1) | 18 / 32 (56.3) | 41 / 45 (91.1) | 6 / 7 (85.7) |
| Sensitivity | 0.67 (0.47, 0.83) | 0.60 (0.53, 0.67) | 0.55 (0.46, 0.64) | 0.58 (0.47, 0.69) | 0.49 (0.34, 0.64) | 0.33 (0.13, 0.59) | 0.61 (0.45, 0.71) | 0.83 (0.36, 1.00) |
| Specificity | 1.00 (0.16, 1.00) | 0.83 (0.69, 0.93) | 0.83 (0.65, 0.94) | 0.83 (0.65, 0.94) | 0.76 (0.53, 0.92) | 0.71 (0.42, 0.92) | 0.75 (0.19, 0.91) | 0.00 (0.00, 0.97) |
| PPV<sub>obs</sub><sup>d</sup> | 1.00 (0.83, 1.00) | 0.94 (0.88, 0.97) | 0.93 (0.84, 0.98) | 0.91 (0.80, 0.97) | 0.82 (0.63, 0.94) | 0.60 (0.26, 0.88) | 0.96 (0.80, 1.00) | 0.83 (0.36, 1.00) |
| NPV<sub>obs</sub><sup>d</sup> | 0.17 (0.02, 0.48) | 0.33 (0.24, 0.43) | 0.32 (0.22, 0.44) | 0.41 (0.29, 0.54) | 0.40 (0.25, 0.57) | 0.45 (0.24, 0.68) | 0.16 (0.03, 0.41) | 0.00 (0.00, 0.97) |
| PPV<sub>50</sub><sup>d</sup> | 1.00 (0.71, 1.00) | 0.78 (0.68, 0.87) | 0.77 (0.63, 0.87) | 0.78 (0.63, 0.89) | 0.67 (0.46, 0.85) | 0.54 (0.21, 0.84) | 0.71 (0.46, 0.85) | 0.45 (0.10, 0.85) |
| NPV<sub>50</sub><sup>d</sup> | 0.75 (0.52, 0.91) | 0.68 (0.59, 0.75) | 0.65 (0.55, 0.74) | 0.67 (0.55, 0.77) | 0.60 (0.44, 0.74) | 0.52 (0.30, 0.73) | 0.66 (0.45, 0.83) | 0.00 (0.00, 1.00) |

496<sup>a</sup> Mass spectrometry approach taken does not discriminate between tetracycline and doxycycline.497<sup>b</sup> Concentration at or above the lowest minimal inhibitory concentration (see methods) and prior descriptions<sup>51</sup>498<sup>c</sup> Reference standard to define positive cholera cases is culture or qPCR (*tcpA*). Approach independent of phage exposure.499<sup>d</sup> Subscript indicates the proportion of samples that were gold-standard positive in the given group for the calculation of each predictive value, with "obs"=observed and "50"=50%. For example, PPV<sub>50</sub> or standardized positive predictive value indicates the positive predictive value of RDT in each group if 50% of that group was gold-standard positive for cholera.502<sup>e</sup> Patients who met the following criteria had subsequent mass spectrometry: (i) culture positive for Vc, (ii) phage (ICP1,2,3) positive by PCR among culture-negative samples, or (iii) Vc positive by PCR among a random 10% of samples negative by culture for Vc and by PCR for phage (see methods).

**Table S3.** Among *Vc* positive samples, impact of antibiotic exposure and resistance on RDT positivity (phage removed)^a^.

| Condition | RDT+ | RDT-<br>(false negatives) | OR (CI) | P value<sup>b</sup> |
| --- | --- | --- | --- | --- |
| Ciprofloxacin (LC-MS/MS qual detection)<sup>c</sup> |  |  |  |  |
| Exposed | 30% (47) | 22% (34) | 0.705 (0.365-1.352) | 0.3234 |
| Not exposed | 32% (49) | 16% (25) |  |  |
| Azithromycin (LC-MS/MS qual detection) |  |  |  |  |
| Exposed | 10% (15) | 10% (16) | 0.498 (0.223-1.105) | 0.099 |
| Not exposed | 52% (81) | 28% (43) |  |  |
| Doxycycline (LC-MS/MS qual detection)<sup>c</sup> |  |  |  |  |
| Exposed | 12% (19) | 7% (11) | 1.077 (0.478-2.523) | 1.000 |
| Not exposed | 50% (77) | 31% (48) |  |  |
| No azithromycin detected by qualitative LC-MS/MS<sup>d</sup> |  |  |  |  |
| Isolate AZI resistant | 25% (28) | 14% (16) | 0.750 (0.337-1.679) | 0.54 |
| Isolate AZI sensitive | 43% (49) | 18% (21) |  |  |
| Azithromycin detected by qualitative LC-MS/MS<sup>d</sup> |  |  |  |  |
| Isolate AZI resistant | 29% (8) | 21% (6) | 1.333 (0.300 6.100) | 1.000 |
| Isolate AZI sensitive | 25% (7) | 25% (7) |  |  |

<sup>a</sup> Criteria to define cholera patients for subsequent mass spectrometry were (i) culture positive for Vc, (ii) phage (ICP1,2,3) positive by PCR among culture-negative samples, or (iii) Vc positive by PCR among a random 10% of samples negative by culture for Vc and by PCR for phage (see methods). Analyses removed samples with ICP1, 2, 3 detected. Those samples without RDT performed were excluded.
<sup>b</sup> Fisher's exact test
<sup>c</sup> Insufficient rates of resistance/sensitive strains for stratified analyses
<sup>d</sup> Among cholera patients in which the culture was positive and an isolate was obtained for AMR testing.

**Table S4.**
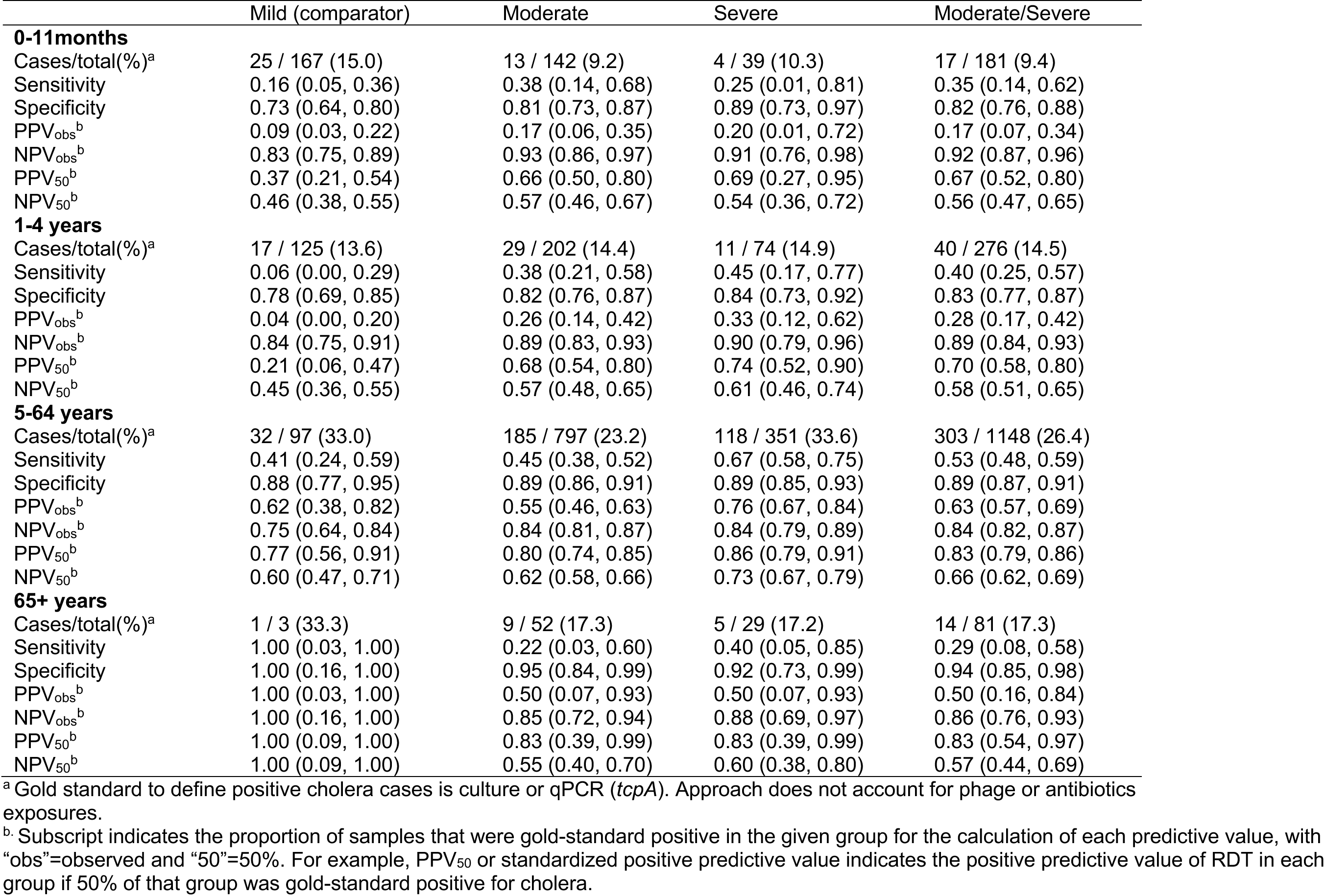
RDT performance by severity and age group with culture or qPCR (*tcpA*) as the reference standard.

|  | Mild (comparator) | Moderate | Severe | Moderate/Severe |
| --- | --- | --- | --- | --- |
| **0-11months** |  |  |  |  |
| Cases/total(%)<sup>a</sup> | 25 / 167 (15.0) | 13 / 142 (9.2) | 4 / 39 (10.3) | 17 / 181 (9.4) |
| Sensitivity | 0.16 (0.05, 0.36) | 0.38 (0.14, 0.68) | 0.25 (0.01, 0.81) | 0.35 (0.14, 0.62) |
| Specificity | 0.73 (0.64, 0.80) | 0.81 (0.73, 0.87) | 0.89 (0.73, 0.97) | 0.82 (0.76, 0.88) |
| PPV<sub>obs</sub><sup>b</sup> | 0.09 (0.03, 0.22) | 0.17 (0.06, 0.35) | 0.20 (0.01, 0.72) | 0.17 (0.07, 0.34) |
| NPV<sub>obs</sub><sup>b</sup> | 0.83 (0.75, 0.89) | 0.93 (0.86, 0.97) | 0.91 (0.76, 0.98) | 0.92 (0.87, 0.96) |
| PPV<sub>50</sub><sup>b</sup> | 0.37 (0.21, 0.54) | 0.66 (0.50, 0.80) | 0.69 (0.27, 0.95) | 0.67 (0.52, 0.80) |
| NPV<sub>50</sub><sup>b</sup> | 0.46 (0.38, 0.55) | 0.57 (0.46, 0.67) | 0.54 (0.36, 0.72) | 0.56 (0.47, 0.65) |
| **1-4 years** |  |  |  |  |
| Cases/total(%)<sup>a</sup> | 17 / 125 (13.6) | 29 / 202 (14.4) | 11 / 74 (14.9) | 40 / 276 (14.5) |
| Sensitivity | 0.06 (0.00, 0.29) | 0.38 (0.21, 0.58) | 0.45 (0.17, 0.77) | 0.40 (0.25, 0.57) |
| Specificity | 0.78 (0.69, 0.85) | 0.82 (0.76, 0.87) | 0.84 (0.73, 0.92) | 0.83 (0.77, 0.87) |
| PPV<sub>obs</sub><sup>b</sup> | 0.04 (0.00, 0.20) | 0.26 (0.14, 0.42) | 0.33 (0.12, 0.62) | 0.28 (0.17, 0.42) |
| NPV<sub>obs</sub><sup>b</sup> | 0.84 (0.75, 0.91) | 0.89 (0.83, 0.93) | 0.90 (0.79, 0.96) | 0.89 (0.84, 0.93) |
| PPV<sub>50</sub><sup>b</sup> | 0.21 (0.06, 0.47) | 0.68 (0.54, 0.80) | 0.74 (0.52, 0.90) | 0.70 (0.58, 0.80) |
| NPV<sub>50</sub><sup>b</sup> | 0.45 (0.36, 0.55) | 0.57 (0.48, 0.65) | 0.61 (0.46, 0.74) | 0.58 (0.51, 0.65) |
| **5-64 years** |  |  |  |  |
| Cases/total(%)<sup>a</sup> | 32 / 97 (33.0) | 185 / 797 (23.2) | 118 / 351 (33.6) | 303 / 1148 (26.4) |
| Sensitivity | 0.41 (0.24, 0.59) | 0.45 (0.38, 0.52) | 0.67 (0.58, 0.75) | 0.53 (0.48, 0.59) |
| Specificity | 0.88 (0.77, 0.95) | 0.89 (0.86, 0.91) | 0.89 (0.85, 0.93) | 0.89 (0.87, 0.91) |
| PPV<sub>obs</sub><sup>b</sup> | 0.62 (0.38, 0.82) | 0.55 (0.46, 0.63) | 0.76 (0.67, 0.84) | 0.63 (0.57, 0.69) |
| NPV<sub>obs</sub><sup>b</sup> | 0.75 (0.64, 0.84) | 0.84 (0.81, 0.87) | 0.84 (0.79, 0.89) | 0.84 (0.82, 0.87) |
| PPV<sub>50</sub><sup>b</sup> | 0.77 (0.56, 0.91) | 0.80 (0.74, 0.85) | 0.86 (0.79, 0.91) | 0.83 (0.79, 0.86) |
| NPV<sub>50</sub><sup>b</sup> | 0.60 (0.47, 0.71) | 0.62 (0.58, 0.66) | 0.73 (0.67, 0.79) | 0.66 (0.62, 0.69) |
| **65+ years** |  |  |  |  |
| Cases/total(%)<sup>a</sup> | 1 / 3 (33.3) | 9 / 52 (17.3) | 5 / 29 (17.2) | 14 / 81 (17.3) |
| Sensitivity | 1.00 (0.03, 1.00) | 0.22 (0.03, 0.60) | 0.40 (0.05, 0.85) | 0.29 (0.08, 0.58) |
| Specificity | 1.00 (0.16, 1.00) | 0.95 (0.84, 0.99) | 0.92 (0.73, 0.99) | 0.94 (0.85, 0.98) |
| PPV<sub>obs</sub><sup>b</sup> | 1.00 (0.03, 1.00) | 0.50 (0.07, 0.93) | 0.50 (0.07, 0.93) | 0.50 (0.16, 0.84) |
| NPV<sub>obs</sub><sup>b</sup> | 1.00 (0.16, 1.00) | 0.85 (0.72, 0.94) | 0.88 (0.69, 0.97) | 0.86 (0.76, 0.93) |
| PPV<sub>50</sub><sup>b</sup> | 1.00 (0.09, 1.00) | 0.83 (0.39, 0.99) | 0.83 (0.39, 0.99) | 0.83 (0.54, 0.97) |
| NPV<sub>50</sub><sup>b</sup> | 1.00 (0.09, 1.00) | 0.55 (0.40, 0.70) | 0.60 (0.38, 0.80) | 0.57 (0.44, 0.69) |

<sup>a</sup> Gold standard to define positive cholera cases is culture or qPCR (*tcpA*). Approach does not account for phage or antibiotics exposures.
<sup>b</sup>. Subscript indicates the proportion of samples that were gold-standard positive in the given group for the calculation of each predictive value, with "obs"=observed and "50"=50%. For example, PPV<sub>50</sub> or standardized positive predictive value indicates the positive predictive value of RDT in each group if 50% of that group was gold-standard positive for cholera.

